# Daily tenofovir disoproxil fumarate/emtricitabine and hydroxychloroquine for pre-exposure prophylaxis of COVID-19: a double-blind placebo controlled randomized trial in healthcare workers

**DOI:** 10.1101/2022.03.02.22271710

**Authors:** R Polo, X García-Albéniz, C Terán, M Morales, D Rial-Crestelo, MA Garcinuño, M García del Toro, C Hita, JL Gómez-Sirvent, L Buzón, A Díaz de Santiago, JL Pérez Arellano, J Sanz, P Bachiller, E Martínez Alfaro, V Díaz-Brito, M Masiá, A Hernández-Torres, J Guerra, J Santos, P Arazo, L Muñoz, JR Arribas, P Martínez de Salazar, S Moreno, MA Hernán, J Del Amo, EPICOS

**Affiliations:** Division for HIV, STI, Viral Hepatitis and TB Control. Ministry of Health, Madrid, Spain; CAUSALab, Harvard T.H. Chan School of Public Health, Boston, Massachusetts, USA; RTI Health Solutions, Barcelona, Spain; Facultad de Medicina Universidad Mayor, Real y Pontificia de San Francisco Xavier de Chuquisaca - Hospital Santa Bárbara, Sucre, Bolivia; Hospital Militar Dr. Carlos Arvelo, Caracas, Venezuela; Hospital Doce de Octubre, Madrid, Spain; CIBER de Enfermedades Infecciosas. Institute of Health Carlos III, Madrid, Spain; Hospital Nuestra Señora de Sonsoles, Ávila, Spain; Hospital General de Valencia, Spain; Hospital de Torrejón, Madrid, Spain; Hospital Universitario de Canarias, Santa Cruz de Tenerife, Spain; Hospital Universitario de Burgos, Burgos, Spain; Hospital Universitario Puerta de Hierro, Madrid, Spain; Hospital Universitario Insular de Gran Canaria, Gran Canaria, Spain; Hospital Universitario de la Princesa, Madrid, Spain; Hospital General de Segovia. Segovia, Spain; Complejo Hospitalario Universitario de Albacete, Albacete, Spain; Parc Sanitari Sant Joan de Déu, Barcelona, Spain; Hospital General Universitario de Elche, Alicante, Spain; Hospital Clínico Universitario Virgen de la Arrixaca, Murcia, Spain; Hospital Universitario de León, León, Spain; Hospital Universitario Virgen de la Victoria de Málaga, Málaga, Spain; Hospital Universitario Miguel Servet. Zaragoza, Spain; Complejo Hospitalario Universitario de Granada, Granada, Spain; Hospital Universitario La Paz, IdiPAZ, Madrid, Spain; Center for Communicable Disease Dynamics, Department of Epidemiology. Harvard T.H. Chan School of Public Health, Boston, Massachsetts, USA; Hospital Universitario Ramón y Cajal, Madrid, Spain; Departments of Epidemiology and Biostatistics, Harvard T.H. Chan School of Public Health, Boston, Massachusetts, USA

## Abstract

**Objective:** To assess the effect of hydroxychloroquine (HCQ), Tenofovir disoproxil fumarate/Emtricitabine (TDF/FTC), and their combination as pre-exposure prophylaxis on the risk of symptomatic COVID-19.

**Methods:** EPICOS is a double-blind, placebo-controlled randomized trial conducted in 51 hospitals in Spain, Bolivia, and Venezuela. Healthcare workers with negative SARS-CoV-2 IgM/IgG test were randomly assigned to: daily TDF/FTC plus HCQ for 12 weeks, TDF/FTC plus HCQ placebo, HCQ plus TDF/FTC placebo and TDF/FTC placebo plus HCQ placebo. The primary outcome was laboratory-confirmed, symptomatic COVID-19. We also studied any (symptomatic or asymptomatic) COVID-19 infection. We compared group-specific 14-week risks via differences and ratios with 95% confidence intervals (CI).

**Results:** Of 1002 individuals screened, 926 (92.4%) were eligible; 64.2% recruited in Spain, 22.3% in Bolivia, and 13.6% in Venezuela. Median age was 38 years (range 18 - 68), 62.5% were female, 62.3% worked at inpatient care, and comorbidities were rare. Compared with the placebo group, 14-week risk ratios (95% CI) of symptomatic COVID-19 were 0.39 (0.00, 1.98) for TDF+HCQ, 0.34 (0.00, 2.06) for TDF, and 0.49 (0.00, 2.29) for HCQ. Corresponding risk ratios of any COVID-19 were 0.51 (0.21, 1.00) for TDF+HCQ, 0.81 (0.44, 1.49) for TDF, and 0.73 (0.41, 1.38) for HCQ. Adverse events were generally mild.

**Conclusion:** A beneficial effect of TDF/FTC and HCQ, alone or in combination, as pre-exposure prophylaxis for COVID-19 cannot be ruled out but effect estimates are imprecise because the target sample size was not met. These findings support launching randomized trials of TDF/FTC for the early treatment of COVID-19.

## Introduction

The repurposing of drugs as prophylactic agents against COVID-19 started early in the pandemic (1). Based largely on in-vitro evidence suggesting hydroxychloroquine (HCQ) may block viral and cell fusion, randomized trials of HCQ as pre-exposure prophylaxis were among the earliest to be launched (1-5). However, these trials were small and resulted in imprecise effect estimates (2-5). Tenofovir disoproxil fumarate (TDF) was another appropriate candidate to test in clinical trials based on epidemiological data suggesting a protective effect against severe COVID-19 (6-7), in-vitro studies reporting inhibition of the SARS-CoV-2 RNA-dependent RNA-polymerase (8-13) and its high bioavailability in many tissues (14-16). However, no randomized trials of TDF for pre-exposure prophylaxis have been completed.

Both hydroxychloroquine and TDF are generic drugs widely prescribed worldwide. HCQ has been used as treatment and prophylaxis of malaria. TDF, in combination with emtricitabine (FTC), has been used for the treatment and prophylaxis of HIV infection. Both drugs have a proven, and well documented, safety record (17-19). Despite their potential for massive use against COVID-19, these safe and inexpensive drugs have not been studied in randomized trials (TDF) or the randomized trials have been relatively small (HCQ).

We carried out a double-blind placebo-controlled randomized trial to assess the effect of daily HCQ or TDF/FTC, and of their combination, during 12 weeks as pre-exposure prophylaxis against COVID-19 in healthcare workers. Here we report estimates of effect and safety.

## Methods

EPICOS (NCT04334928, EudraCT number 2020-001385-11) was a multicenter, double-blind, placebo-controlled randomized trial to study the effect of TDF/FTC and HCQ as pre-exposure prophylaxis for symptomatic COVID-19 among healthcare workers in Spain, Bolivia and Venezuela. The trial was designed to recruit 4000 individuals, but the start of the vaccination campaign and other factors limited recruitment to 907 participants.

Recruitment into the trial was actively promoted in Spain through regional health authorities, the Ministry of Health and in Latin America through Esther (Ensemble de Solidarité Thérapeutique Hospitalière En Reseau). Healthcare workers were approached individually and collectively through promotional in-hospital sessions, mailings and hospital-wide advertisements. A mobile phone app was developed for electronic monitoring, weekly reminders of adherence, and side-effects reporting.

### Eligibility criteria

Healthcare workers aged 18 to 70 years were eligible if they did not have a prior diagnosis of SARS-CoV-2 infection, did not have symptoms compatible with SARS-CoV-2 infection, had a negative IgM/IgG test for SARS-CoV-2, had a negative HIV and (for women) pregnancy tests, a normal electrocardiogram (ECG), and no history of prolongation of the QT interval, maculopathy, impaired renal function, or immunosuppressive or hematologic conditions. Because women comprised the majority of healthcare workers, we ensured 40% of individuals screened for eligibility were males. Recruitment started in April 2020 in Spain, October 2020 in Bolivia, and March 2021 in Venezuela (Supplementary Figure 1). The study ended on May 30, 2021.

### Treatment strategies and assignment

Eligible individuals were randomly assigned to one of four treatment groups: TDF/FTC plus HCQ, TDF/FTC plus HCQ placebo, HCQ plus TDF/FTC placebo and TDF/FTC placebo plus HCQ placebo. TDF/FTC was administered as a single pill with 245mg of TDF and 200 mg of FTC once daily). HCQ was administered as 200 mg once daily, the minimum dose to reach adequate tissue distribution (20). Participants received treatment for 12 weeks (or until a SARS-CoV-2 infection was diagnosed), irrespective of symptoms, or the administrative end of the study, whichever occurred first. Investigators, participants, and the data analyst were unaware of treatment assignment.

### Outcomes

The primary outcome was symptomatic COVID-19, defined as the presence of SARS-CoV-2 infection confirmed by a polymerase chain reaction (PCR) test plus any of the following symptoms: general malaise, fever, cough, joint pain, or breathing difficulty. PCR-confirmed asymptomatic SARS-CoV-2 infection was a secondary outcome. Other secondary outcomes (severity and duration of symptoms) could not be studied. We also studied the outcome “any (symptomatic or asymptomatic) COVID-19 infection”, which had not been pre-specified in the study protocol.

Adverse events were ascertained in each monthly visit and weekly through app reminders. Adverse events were classified as mild (easily tolerated), moderate (interference with normal activities), or severe (incapacitating, with inability to perform normal activities). Regardless of severity, adverse events were classified as serious if they required hospitalization, prolonged an existing hospitalization, or led to major or permanent disability.

### Follow-up

Participants attended three monthly visits after randomization. In each visit, they were evaluated for the presence of adverse events, received standard laboratory tests, IgM/IgG antibody test for SARS-CoV-2, and an ECG if necessary. A PCR test was performed if the IgM/IgG antibody test was positive or if symptoms were present. A fourth monthly visit was scheduled for the evaluation of adverse events only.

### Statistical Analysis

We used the Kaplan-Meier estimator to obtain the cumulative risk for each outcome over 14 weeks of follow-up in each treatment group (over 95% of participants had attended their third monthly visit by 14 weeks after randomization). We compared the group-specific risks via differences and ratios with the placebo only group as the reference. In post-hoc analyses, we compared the risk in the two groups containing HCQ with the two groups not containing HCQ, and in the two groups containing TDF/FTC with the two groups not containing TDF/FTC. We calculated 95% confidence intervals using the percentile bootstrap method with 500 repetitions. In sensitivity analyses, we used a Cox proportional hazards model to estimate the hazard ratio of the two outcomes.

This study was approved by the institutional review boards of University Hospital de La Princesa, Madrid, Spain, Servicio Departamental de Salud de Chuquisaca in Bolivia, and Instituto Nacional de Higiene “Rafael Rangel” in Venezuela. An independent medical monitor and a data safety monitoring board provided oversight of safety and efficacy.

## Results

Of 1002 individuals screened for eligibility, 926 (92.4%) were eligible. The main reason for ineligibility was a previous COVID-19 diagnosis or compatible symptoms (Figure 1). Nineteen individuals withdrew or were lost to follow-up before treatment assignment. Of 907 randomized individuals, 220 were assigned to the TDF/FTC plus HCQ group (12 did not start treatment), 231 to the TDF/FTC placebo plus HCQ group (7 did not start treatment), 233 to the TDF/FTC plus HCQ placebo group (12 did not start treatment), and 223 to the double placebo group (12 did not start treatment). Of 696 individuals who completed the scheduled follow-up, 668 completed the scheduled treatment as indicated in the protocol. Supplementary Tables 1 and 2 show the reasons for early termination of treatment and incomplete follow-up, respectively, by treatment group.

**Figure 1.**
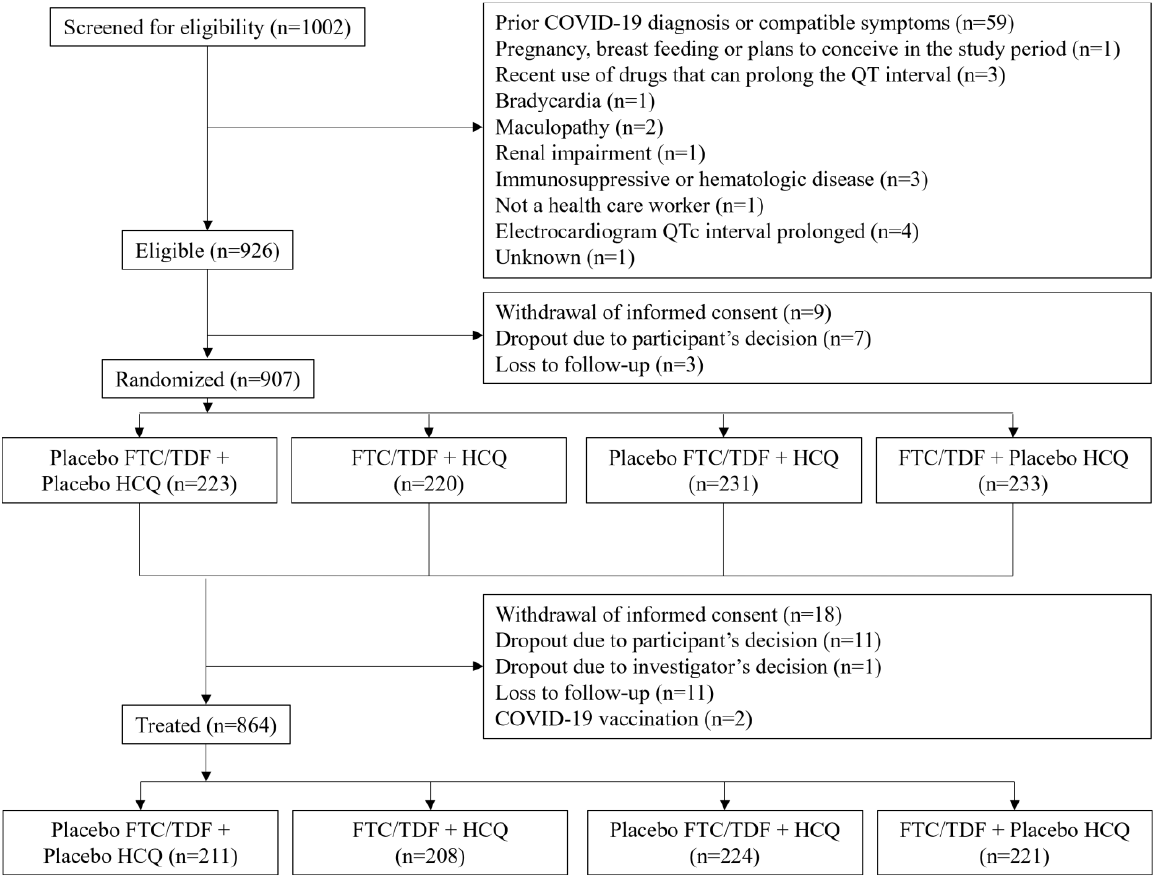
Flowchart of participants, EPICOS randomized trial TDF/FTC: Tenofovir/emtricitabine; HCQ: hydroxychloroquine

Baseline characteristics of the 907 participants are summarized in Table 1 and Supplementary Table 3. Their median age was 38 years (range 18 to 68 years) and 62.5% were female. Most participants worked at inpatient care facilities (62.3%) and the most frequent occupation was physician (30.5%). Comorbidities were rare. By country, 64.2% of participants were recruited in Spain, 22.3% in Bolivia, and 13.6% in Venezuela.

**Table 1.** Baseline characteristics of 907 participants, EPICOS randomized trial

| Characteristic | TDF/FTC<br>+ HCQ<br>(n=220) | TDF/FTC<br>(n=233) | HCQ<br>(n=231) | Placebo<br>(n=223) |
| --- | --- | --- | --- | --- |
| Sex, n (%) |  |  |  |  |
| Male | 85 (38.6) | 93 (39.9) | 82 (35.5) | 80 (35.9) |
| Female | 135 (61.4) | 140 (60.1) | 149 (64.5) | 143 (64.1) |
| Age in years,<br>median (range) | 38.0<br>(18.0, 65.0) | 39.0<br>(18.0, 68.0) | 38.0<br>(18.0, 65.0) | 38.0<br>(18.0, 65.0) |
| Occupation, n (%) |  |  |  |  |
| Physician | 71 (32.3%) | 68 (29.2%) | 74 (32.0%) | 66 (29.6%) |
| Nurse | 63 (28.6%) | 77 (33.0%) | 67 (29.0%) | 72 (32.3%) |
| Medical student on<br>clinical rotation | 59 (26.8) | 58 (24.9) | 59 (25.5) | 53 (23.8) |
| Other, with direct<br>patient contact | 13 (5.9%) | 13 (5.6%) | 11 (4.8%) | 11 (4.9%) |
| Other, without direct<br>patient contact | 13 (5.9%) | 10 (4.3%) | 15 (6.5%) | 18 (8.1%) |
| Unknown | 1 (0.5%) | 7 (3.0%) | 5 (2.2%) | 3 (1.3%) |
| Comorbidities, n (%) |  |  |  |  |
| Cardiac disease | 3 (1.4) | 0 | 1 (0.4) | 2 (0.9) |
| Hypertension | 17 (7.7) | 15 (6.4) | 4 (1.7) | 19 (8.5) |
| Pulmonary disease | 0 | 0 | 0 | 0 |
| Asthma | 17 (7.7) | 8 (3.4) | 20 (8.7) | 9 (4.0) |
| Neoplasia | 4 (1.8) | 4 (1.7) | 2 (0.9) | 1 (0.4) |
| Diabetes | 4 (1.8) | 3 (1.3) | 1 (0.4) | 3 (1.3) |
| Autoimmune<br>Disease | 5 (2.3) | 7 (3.0) | 4 (1.7) | 2 (0.9) |
| Country, n (%) |  |  |  |  |
| Spain | 139 (63.2%) | 151 (64.8%) | 148 (64.1%) | 144 (64.6%) |
| Venezuela | 31 (14.1%) | 31 (13.3%) | 32 (13.9%) | 29 (13.0%) |
| Bolivia | 50 (22.7%) | 51 (21.9%) | 51 (22.1%) | 50 (22.4%) |

<sup>a</sup>This category includes, biochemists, hospital chaplains, clinical data managers, dentists, psychologists, janitors, human resources personnel, and social workers.

Figure 2A shows the cumulative risk of symptomatic COVID-19 by treatment group. There were 14 cases: 3 in each group with active treatment and 5 in the placebo only group. All cases had mild symptoms that did not require hospitalization (Supplementary Table 4). Compared with the placebo only group, the 14-week risk ratio (95% CI) of symptomatic COVID-19 was 0.39 (0.00, 1.98) for TDF+HCQ, 0.34 (0.00, 2.06) for TDF, and 0.49 (0.00, 2.29) for HCQ (Table 2).

**Table 2.**
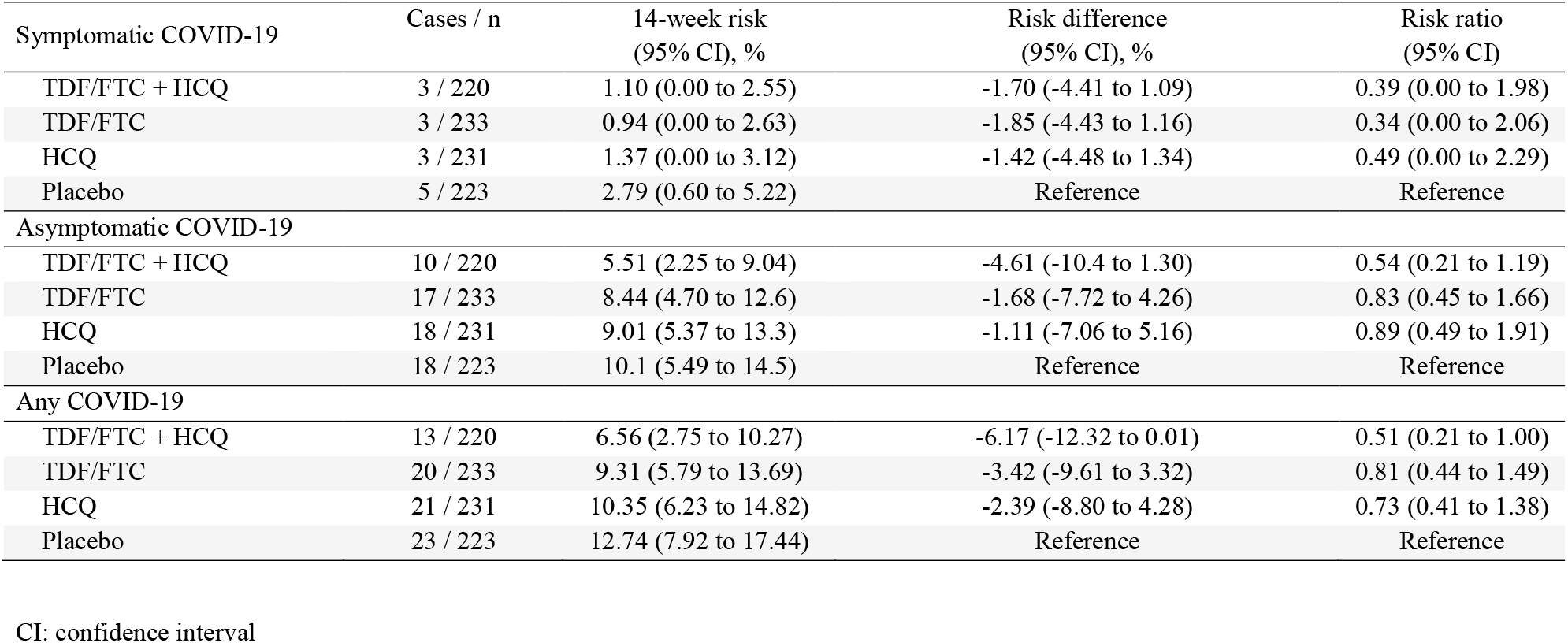
Estimated 14-week risks of symptomatic, asymptomatic, and any COVID-19 diagnosis by treatment group, EPICOS randomized trial

**Figure 2.**
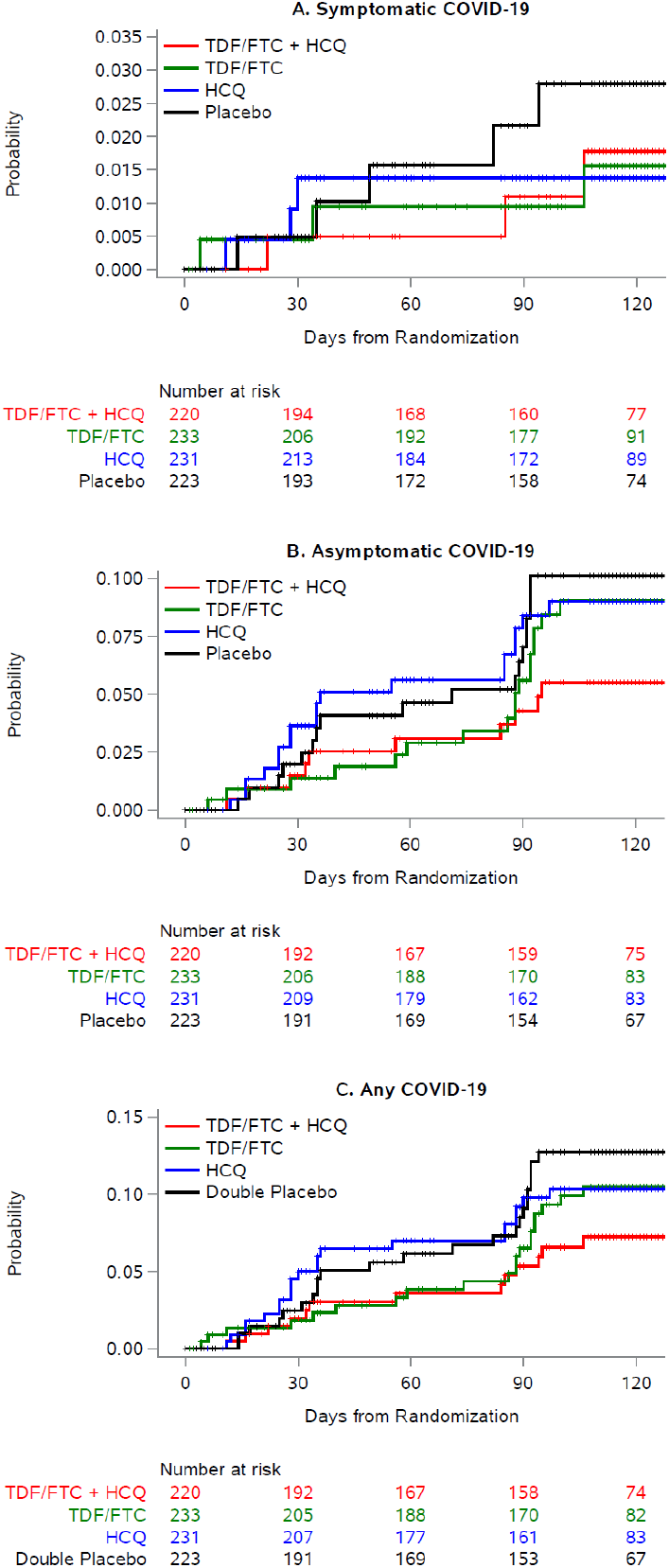
Cumulative risk of symptomatic and asymptomatic COVID-19 by treatment group, EPICOS randomized trial

The 14-week risk ratio (95% CI) of symptomatic COVID-19 was 0.68 (0.10, 2.04) for the groups assigned to HCQ compared with the two groups not assigned to HCQ (Supplementary Table 5 and Supplementary Figure 2), and 0.49 (0.09, 1.70) for the groups assigned to TDF/FTC compared with the two groups not assigned to TDF/FTC (Supplementary Table 6 and Supplementary Figure 3).

Figure 2B shows the cumulative risk of asymptomatic COVID-19 by treatment group. There were 63 cases: 10 in the TDF/FTC + HC group, 17 in the TDF/FTC group, 18 in the HCQ group, and 17 in the placebo only group. Compared with the placebo only group, the 14-week risk ratio (95% CI) of symptomatic COVID-19 was 0.54 (0.21, 1.19) for TDF+HCQ, 0.83 (0.45, 1.66) for TDF, and 0.89 (0.49, 1.91) for HCQ (Table 2).

The 14-week risk ratio (95% CI) of asymptomatic COVID-19 was 0.79 (0.47, 1.33) for the groups assigned to HCQ compared with the groups not assigned to HCQ (Supplementary Table 5 and Supplementary Figure 2), and 0.74 (0.43, 1.21) for the groups assigned to TDF/FTC compared with the groups not assigned to TDF/FTC (Supplementary Table 6 and Supplementary Figure 3).

Figure 2C shows the cumulative risk of any COVID-19 diagnosis by treatment group. There were 77 cases: 13 in the TDF/FTC + HC group, 20 in the TDF/FTC group, 21 in the HCQ group, and 23 in the placebo only group. Compared with the placebo only group, the 14-week risk ratio (95% CI) of any COVID-19 diagnosis was 0.51 (0.21, 1.00) for TDF+HCQ, 0.81 (0.44, 1.49) for TDF, and 0.73 (0.41, 1.38) for HCQ (Table 2).

The 14-week risk ratio (95% CI) of any COVID-19 diagnosis was 0.78 (0.49, 1.23) for the groups assigned to HCQ compared with the groups not assigned to HCQ (Supplementary Table 5 and Supplementary Figure 2), and 0.70 (0.43, 1.10) for the groups assigned to TDF/FTC compared with the groups not assigned to TDF/FTC (Supplementary Table 6 and Supplementary Figure 3).

The corresponding hazard ratios were similar (Supplementary Table 7).

The proportion of individuals with adverse events ranged between 21.1% in the placebo group and 31.3% in the TDF/FTC group. Most were mild and of gastrointestinal nature (Table 3). There were 5 serious adverse events: 4 in the placebo only group (hospital admission due to a bleeding uterine myoma, hospital admission due to smoke inhalation from a workplace fire, an episode of dizziness and bradypsiquia, and an episode of jaundice and vomiting) and 1 in the TDF/FTC +HCQ group (retinal detachment). None of the serious adverse events were linked to the study drugs.

**Table 3.** Frequency of adverse events^*^ by treatment group, EPICOS randomized trial

|  | TDF/FTC + HCQ (n=220) | TDF/FTC (n=233) | HCQ (n=231) | Placebo (n=223) |
| --- | --- | --- | --- | --- |
| Severity of adverse event |  |  |  |  |
| Mild | 78 (35.5%) | 77 (33.0%) | 63 (27.3%) | 63 (28.3%) |
| Moderate | 37 (16.8%) | 33 (14.2%) | 36 (15.6%) | 29 (13.0%) |
| Severe | 1 (0.5%) | 1 (0.4%) | 1 (0.4%) | 2 (0.9%) |
| Adverse event classified as serious | 1 (0.5%) | 0 | 0 | 4 (1.8%) |
| Adverse event classified as related to study drug | 49 (22.3%) | 51 (21.9%) | 46 (19.9%) | 37 (16.6%) |
| Impact of adverse event on study treatment |  |  |  |  |
| Treatment was interrupted | 28 (12.7%) | 27 (11.6%) | 14 (6.1%) | 19 (8.5%) |
| Treatment was delayed | 4 (1.8%) | 4 (1.7%) | 7 (3.0%) | 3 (1.3%) |
| Concomitant treatment was prescribed | 23 (10.5%) | 26 (11.2%) | 23 (10.0%) | 21 (9.4%) |
| Adverse events by system organ class<sup>a</sup> |  |  |  |  |
| Gastrointestinal disorders | 68 (30.9%) | 73 (31.3%) | 56 (24.2%) | 47 (21.1%) |
| Blood and lymphatic system disorders | 1 (0.5%) | 0 | 0 | 1 (0.4%) |
| Cardiac disorders | 1 (0.5%) | 2 (0.9%) | 1 (0.4%) | 3 (1.3%) |
| Ear and labyrinth disorders | 1 (0.5%) | 2 (0.9%) | 0 | 3 (1.3%) |
| Eye disorder | 3 (1.4%) | 1 (0.4%) | 2 (0.9%) | 4 (1.8%) |
| General disorders | 11 (5.0%) | 17 (7.3%) | 9 (3.9%) | 10 (4.5%) |
| Immune system disorder | 0 | 1 (0.4%) | 0 | 0 |
| Infections | 4 (1.8%) | 0 | 5 (2.2%) | 3 (1.3%) |
| Injuries | 2 (0.9%) | 0 | 1 (0.4%) | 2 (0.9%) |
| Investigations | 2 (0.9%) | 6 (2.6%) | 3 (1.3%) | 3 (1.3%) |
| Metabolism and nutrition disorders | 2 (0.9%) | 2 (0.9%) | 1 (0.4%) | 1 (0.4%) |
| Musculoskeletal / connective tissue disorders | 9 (4.1%) | 9 (3.9%) | 6 (2.6%) | 6 (2.7%) |
| Nervous system disorders | 22 (10.0%) | 31 (13.3%) | 26 (11.3%) | 19 (8.5%) |
| Psychiatric disorders | 3 (1.4%) | 3 (1.3%) | 4 (1.7%) | 8 (3.6%) |
| Renal and urinary disorders | 0 | 1 (0.4%) | 0 | 1 (0.4%) |
| Reproductive system disorder | 1 (0.5%) | 0 | 1 (0.4%) | 1 (0.4%) |
| Respiratory disorders | 1 (0.5%) | 3 (1.3%) | 3 (1.3%) | 2 (0.9%) |
| Skin disorders | 14 (6.4%) | 6 (2.6%) | 6 (2.6%) | 4 (1.8%) |
| Vascular disorders | 0 | 0 | 1 (0.4%) | 3 (1.3%) |

\* more than one adverse event per participant could occur
<sup>a</sup> See supplementary methods for a list of the observed adverse events in each system organ class

## Discussion

In this randomized, double-blind, placebo-controlled clinical trial we evaluated the effect of treatment with HCQ and TDF/FTC, alone or in combination, as pre-exposure prophylaxis for symptomatic COVID-19 among health care workers after 12 weeks of follow-up. However, the trial recruited approximately a quarter of the intended number of participants and thus the effect estimates were imprecise: compared with placebo, the risk of symptomatic COVID-19 was lower in the groups assigned to HCQ or TDF/FTC, but effects between a 2-fold risk increase and perfect protection with HCQ or TDF/FTC were highly compatible with the data. For any (symptomatic or asymptomatic) COVID-19, the risk in the group assigned to combined HCQ plus TDF/FTC was half the risk in the group assigned to placebo only, and effects between a 79% reduction in risk and no reduction in risk were highly compatible with the data.

EPICOS also confirmed HCQ and TDF/FTC were safe, with mostly mild adverse events of gastrointestinal nature. This was expected because, after decades of use, the safety record of low-dose HCQ and of TDF/FTC is well established (17-19). TDF/FTC has been shown to be safe even when used during pregnancy (19).

Several randomized, placebo-controlled clinical trials have studied HCQ, at different doses and with different outcome definitions, as pre-exposure prophylaxis for (mostly non-severe) COVID-19 in health care workers (2,4-5). Like EPICOS, five of these trials could not achieve their intended sample size (4,5), partly because potential participants were averse to receive HCQ after poorly conducted observational studies (later retracted) (21) suggested HCQ was not safe, and the “nonsignificant” findings of small randomized trials for prophylaxis were misinterpreted as lack of a beneficial effect. However, all trials with more than 1000 patients found similar estimates: a COVID-19 hazard ratio of 0.73 (95% CI 0.48-1.09) for HCQ vs. placebo after 12 weeks of follow-up in a trial with 1483 participants (5), a COVID-19 odds ratio of 0.75 (95% CI 0.49-1.15) for HCQ vs. placebo after 29 days follow-up in the HERO-HCQ trial with 1359 participants (4), and relative risk of 0.70 (95% CI 0.44-0.97) for HCQ vs. ascorbic acid after 42 days of follow-up in a cluster randomized trial of 1051 participants (22). When taken altogether with the findings from EPICOS, the evidence cannot rule out the possibility that prophylaxis with HCQ offers a modest protection against COVID-19 (3).

No previous randomized trials had studied TDF/FTC as pre-exposure prophylaxis for COVID-19. However, several observational studies have found a lower risk of COVID-19 diagnosis or of hospitalization among individuals who use TDF/FTC compared with those who do not (6-7,23-25). A study among people with HIV in Spain reported lower risk of COVID-19 hospitalization among individuals treated with TDF/FTC compared with those treated with other antiretrovirals (6-7). However, the estimates were not adjusted for the potentially different clinical characteristics of individuals receiving each treatment. A second study in over 50,000 persons with HIV and adequate virological control, which adjusted for comorbidities and other factors, also found a lower risk ratio of COVID-19 hospitalization for TDF/FTC compared with TAF/FTC. Adjusted and unadjusted estimates were similar (24).

A lower risk of COVID-19 hospitalization or death was also found among HIV-positive individuals who used TDF/FTC for HIV treatment in South Africa (23) and among individuals who used TDF for the treatment of hepatitis B infection (25). Also, in a study of ferrets infected with SARS-CoV-2, the group treated with TDF/FTC group showed a reduction in clinical scores and a shorter duration of symptoms (26). A small randomized trial in humans with mild COVID-19 found reductions in nasopharyngeal shedding of SARS-CoV-2 4 days and 7 days after initiation of treatment TDF/FTC (27). On the other hand, a recent in-vitro study report could not detect substantial activity of TDF/FTC against SARS-CoV-2 (28).

The findings from the EPICOS trial add to this growing evidence suggesting a beneficial effect of TDF/FTC in early SARS-CoV-2 infection. EPICOS also confirmed TDF/FTC is safe, as has been found over two decades of use, even in pregnant women (18-19). However, even if HCQ and TDF/FTC were effective as pre-exposure prophylaxis for COVID-19, the practical implications are currently limited because vaccines are a better approach to prevention when available, at least for the variants studied so far and for immunocompetent persons. The efficacy of vaccines seems to be reduced in immunocompromised patients who are in need of other prophylactic strategies (29).

A timelier question is whether HCQ and TDF/FTC could be used for early treatment of COVID-19 in non-hospitalized patients. The question has already been answered for HCQ (30) but not for TDF/FTC, a generic and inexpensive drug combination with the potential for massive worldwide production, and for which the available evidence supports the need for therapeutic trials.

In summary, this randomized, double-blind, placebo-controlled clinical trial in 907 health care workers could not rule out a beneficial effect of HCQ and TDF/FTC, alone or in combination, as pre-exposure prophylaxis for COVID-19. The findings from this trial, combined with previous evidence, support the launching of randomized trials of TDF/FTC for the early treatment of COVID-19.

## Data Availability

All data produced in the present study are available upon reasonable request to the authors

## APPENDIX: EPICOS RESEARCH TEAM

The EPICOS randomized trial was sponsored by the Ministry of Health of Spain.

### EPICOS investigator team

Julia del Amo (Principal Investigator), Ministry of Health, Madrid, Spain

Rosa Polo, Ministry of Health, Madrid, Spain

Santiago Moreno, Hospital Ramón y Cajal, Madrid, Spain

Juan Berenguer, Hospital Gregorio Marañon, Madrid, Spain

Esteban Martínez, Hospital Clinic, Barcelona, Spain

Miguel Hernán, CAUSALab and Department of Epidemiology, Harvard T.H. Chan School of Public Health, Boston, MA, USA

Pablo Martínez de Salazar, Department of Epidemiology, Harvard T.H. Chan School of Public Health, Boston, MA, USA

Xabier García de Albéniz, RTI Health Solutions, Barcelona, Spain

### Collaborators

Marieta Iradier, Fundación Estatal, Salud, Infancia y Bienestar Social

Inma Jarrín, Institute of Health Carlos III, National Center for Epidemiology

### Data safety monitoring board

Javier Zamora, Hospital Ramón y Cajal, Madrid, Spain Antonio Rivero, Hospital Reina Sofía, Córdoba, Spain Clara Menéndez, IS Global, Barcelona, Spain

### Contract Research Organization

Enrique Conde and José Montes, Effice Research, Madrid, Spain

### Participating hospitals

#### Bolivia

- Facultad de Medicina Universidad Mayor, Real y Pontificia de San Francisco Xavier de Chuquisaca -Hospital Santa Bárbara, Bolivia. PI: Carolina Terán. Co-investigators: Bettsy Flores, María Elena Choque, Jhaquelin Peñaranda, Gladys Gorena, Mariluz Herrera, Marcela Farfán, David Moya, Jhonny Camacho, Jovanna Ordoñez, José Mayora, Brayan Farfán

#### Venezuela

- Hospital Militar Dr. Carlos Arvelo, Centro Clínico María Edelmira Araujo, Instituto Falconiano de Emergencias Médicas, Venezuela. PI: Miguel Morales. Co-investigators: Maryelis Benítez, Rosa Bolaños, Jesús Colina

#### Spain

- Hospital Doce de Octubre, Madrid. Co-investigators: Federico Pulido, Rafael Rubio, Otilia Bisbal, María de Lagarde, Cristina Epalza, Cristina Lillo-Díaz, Raúl Martínez
- Hospital Nuestra Señora de Sonsoles, Ávila. Co-investigators: Miguel Sebastián Pedrodomingo, César de la Hoz, Demetrio Sánchez, Ana Cristina Antolí, Carmen Grande, Dulce María Astudillo
- Hospital General de Valencia. PI: Miguel García Deltoro. Co-investigators: Jose Ignacio Chirivella
- Hospital de Torrejón, Madrid. PI: César Hita. Co-investigators: María Carmen Montero, Juan Ruíz.
- Hospital Universitario de Tenerife. Canarias. Co-investigators: Mª Mar Alonso, Mª Remedios Alemán, Ana Mª López, Dácil García, Ricardo Pelazas
- Hospital Universitario de Burgos. Co-investigators: Pablo González Recio
- Hospital Puerta de Hierro, Madrid. Co-investigators: Fernando Martínez-Vera, Alejandro Muñoz, Sara De la Fuente, Ana Muñoz, José Manuel Vázquez
- Hospital Universitario Insular de Gran Canaria. Co-investigators: Cristina Carranza, Michele Hernández, Nieves Jaén, Carmen Lavilla, Elena Pisos, Laura Suárez
- Hospital La Princesa, Madrid. Co-investigators: Ignacio de los Santos, Lucio García-Fraile, Ángela Gutiérrez, Azucena Bautista
- Hospital General de Segovia. Co-investigators: Sara Muñoz, José María Alonso de los Santos, Eva María Ferreira, Ana Carrero
- Complejo Hospitalario Universitario de Albacete. Co-investigators: Fernando Mateos, José Javier Blanch, Julian Eloy Solis García
- Parc Sanitari Sant Joan de Déu. Sant Boi del Llobregat, Barcelona. Co-investigators: Montserrat Sanmartí, Raquel Gómez, Encarna Moreno, María Carmen Álvarez
- Hospital General Universitario de Elche. Co-investigators: Javier García-Abellán. Félix Gutiérrez, Sergio Padilla, Gabriel Estañ
- Hospital Clínico Universitario Virgen de la Arrixaca. Murcia. Co-investigators: Encarnación Moral, Sonia Marín, Aychel Elena Roura, Ana Pareja
- Hospital Universitario de León. Co-investigators: Manuel Martín Regidor, Esperanza Gutiérrez, Luis Jorge Valdivia, Patricia Capón
- Hospital Universitario Virgen de la Victoria de Málaga. Co-investigators: Antonia Domínguez, Antonia Moreno, Luis Ruiz, Rubén Garrido
- Hospital Universitario Miguel Servet. Zaragoza. Co-investigators: Álvaro Cecilio, Rosa Fenoll
- Hospital Clínico San Cecilio (Complejo Hospitalario Universitario de Granada). Co-investigators: José Peregrina, Francisco Anguita, Laura Martín
- Hospital Universitario Ramón y Cajal. Madrid. PI: Fernando Dronda. Co-investigators: Johannes Häemmerle, Clara Crespillo
- Hospital Arnau de Vilanova – Llíria. Valencia. PI: Juan Flores. Co-investigators: Lidia Castellano
- Hospital Clínico Universitario de Valladolid. PI: Carlos Dueñas. Co-investigators: Laura Rodríguez Fernández, Genoveva Zapico
- Hospital La Fe. Valencia. PI: María Tasias. Co-investigators: Pablo Berrocal, Cristina Campo
- Hospital Universitario Virgen Macarena / Instituto de Biomedicina de Sevilla. PI: Jesús Rodríguez Baño. Co-investigators: Ángel Domínguez-Castellano, María José Ríos-Villegas
- Hospital Sant Joan de Déu. Barcelona. PI: Antoni Noguera-Julian. Co-investigators: Clàudia Fortuny, María Ríos-Barnés
- Hospital Universitario de Salamanca. PI: Guillermo Hernández-Pérez. Co-investigators: María Sánchez-Ledesma, Cristina Carbonell
- Complejo Asistencial Universitario de Palencia. PI: Jacinto Sánchez-Navarro. Co-investigators: Cristina Sánchez del Hoyo, Yolanda Morán
- Fundación Jiménez Díaz, Madrid. PI: Alfonso Cabello. Co-investigators: Irene Carrillo, Miguel Górgolas
- Hospital Clínico de Santiago de Compostela. PI: Antonio Antela. Co-investigators: Elena Losada, Maria Jesús Domínguez
- Hospital Universitario de Ferrol. PI: Ana Isabel Mariño. Co-investigators: Sabela Sánchez-Trigo, Silvia Martínez-Varela
- Hospital Infanta Margarita, Córdoba. PI: Eduardo Aguilar. Co-investigators: Jesús González-Lama, Alejandro Plata
- Hospital Reina Sofía Tudela. Navarra. PI: Mª Teresa Rubio. Co-investigators: Marta Marín, Lucía Zardoya
- Hospital Universitari Sagrat Cor -Grupo Quirónsalud. Barcelona. PI: Diego de Mendoza. Co-investigators: Antonio Gutiérrez, Rosa Coll
- Hospital Clínico Universitario Lozano Blesa, Zaragoza. PI: Isabel Sanjoaquín. Co-investigators: Silvia Loscos
- Hospital Virgen de la Luz, Cuenca. PI: MP Geijo. Co-investigators: O Belinchon
- Hospital Reina Sofía, Murcia. PI: Enrique Bernal
- Hospitales Universitarios Rey Juan Carlos, Infanta Elena y General de Villalba, Madrid. PI: Ámbar Deschamps-Perdomo
- Hospital Universitario Príncipe de Asturias, Madrid. PI: José Alberto Arranz
- Hospital Universitario La Paz, Madrid. PI: Alberto Borobia
- Hospital del Mar, Barcelona. PI: Hernando Knobel
- Hospital Universitario Doctor Peset, Valencia. PI: Arturo Artero
- Complejo Hospitalario de Navarra, Pamplona. PI: María Rivero
- Hospital Clínico de Valencia. PI: María José Galindo
- Hospital Universitario de Móstoles. Madrid. PI: Concepción Cepeda
- Hospital San Pedro, Logroño. PI: José Ramón Blanco
- Hospital Clínico San Carlos, Madrid. PI: Vicente Estrada
- Hospital Universitario Araba-Txagorritxu. Vitoria. PI: Ainhoa Lecuona
- Hospital Río Hortega de Valladolid. PI: Julia Gómez
- Hospital Universitario Central de Asturias. PI: Víctor Asensi
- Hospital Universitario Severo Ochoa, Madrid. PI: Miguel Cervero

## Supplementary material

## Supplementary Methods

### Eligibility criteria

#### Inclusion criteria

- Informed consent for participation
- Acknowledgment of understanding of the purpose of the study
- Age 18-70 years
- Health care worker in hospitals and other health facilities in areas at risk of SARS-CoV-2 transmission
- No previous diagnosis of SARS-CoV-2 infection
- Negative rapid test for IgM/IgG for SARS-CoV-2
- Negative pregnancy test during the previous 7 days or >2 years after menopause
- For women of reproductive age and their partners, commitment to use highly effective contraceptive method (double barrier, hormonal contraception) during the study period and at least until 6 months after the last dose of treatment
- Normal ECG

#### Exclusion criteria

- Symptoms suggestive of COVID-19 infection
- HIV infection
- Active hepatitis B infection.
- Estimated glomerular filtration rate (GFR) < 60 ml/min) or on hemodialysis.
- Osteoporosis
- Myasthenia gravis
- Pre-existent maculopathy
- Retinitis pigmentosa
- Bradycardia < 50 bpm
- Weight < 40 kg
- Participant with any immunosuppressive condition or hematological disease.
- Any medication as prophylaxis against SARS-CoV-2 or HIV after March 1, 2020.
- Treatment in the previous month and for >7 days with drugs that may prolong QT interval, including azithromycin, chlorpromazine, cisapride, clarithromycin, domperidone, droperidol, erythromycin, halofantrine, haloperidol, lumefantrine, mefloquine, methadone, pentamidine, procainamide, quinidine, quinine, sotalol, sparfloxacin, thioridazine, amiodarone.
- Pregnancy or plans to conceive during the study period.
- Breastfeeding
- Known allergy to any of the experimental medications or excipients

## Supplementary Results

### Description of observed adverse events by system organ class

- Gastrointestinal disorders: abdominal discomfort/distension/pain, aerophagia, constipation, decreased appetite, diarrhea, dry mouth, dysgeusia, dyspepsia, flatulence, food allergy, gastritis, gastroesophageal reflux disease, hemorrhoids, lip pruritus, nausea, odynophagia, vomiting.
- Blood and lymphatic system disorders: iron deficiency anemia and leukopenia.
- Cardiac disorders: palpitations, tachycardia, left posterior fascicular block and right bundle branch block
- Ear and labyrinth disorders: hypoacusis and vertigo
- Eye disorders: corneal erosion, dry eye, iridocyclitis, photophobia, retinal detachment, blurred vision and loss of visual acuity.
- General disorders: asthenia, chest pain, chills, discomfort, fatigue, feeling hog, malaise, peripheral edema and pyrexia
- Immune system disorder: allergic reaction to arthropod sting
- Infections: bartholinitis, bronchitis, viral conjunctivitis, corneal abscess, ear infection, gastroenteritis, gingivitis, helicobacter infection, oral herpes, pharyngotonsillitis, tinea versicolor and urinary tract infection
- Injuries: skeletal injuries, fall, ligament sprain, smoke inhalation and traffic accident.
- Investigations: abnormal findings in any of the following tests: electrocardiogram, blood creatinine, blood glucose, hemogram, thyroid hormones, body temperature, glomerular filtration rate, liver function, serum ferritin, weight.
- Metabolism and nutrition disorders: hyporexia, increased ferritin and hypertriglyceridemia.
- Musculoskeletal and connective tissue disorders: arthralgia, back pain, bursitis, muscle spasms, muscular weakness, myalgia, neck pain and extremity pain.
- Nervous system disorders: amnesia, burning sensation, dizziness, dysesthesia, dyskinesia, headache, migraine, paresthesia, somnolence and tremor.
- Psychiatric disorders: nightmares, anxiety, bradyphrenia, depressed modo, insomnia, irritability and tearfulness.
- Renal and urinary disorders: nocturia and pollakiuria.
- Reproductive system disorders: dysmenorrhea and increased menstrual bleeding.
- Respiratory disorders: asthmatic crisis, cough, exertional dyspnea, respiratory distress, allergic rhinitis, rhinorrhea and throat irritation.
- Skin disorders: alopecia, allergic dermatitis, erythema, guttate psoriasis, pruritus, rash, skin exfoliation, solar lentigo and urticaria.
- Vascular disorders: hot flush, hypertension, hypertensive crisis and hypotension.

**Supplementary Table 1.**
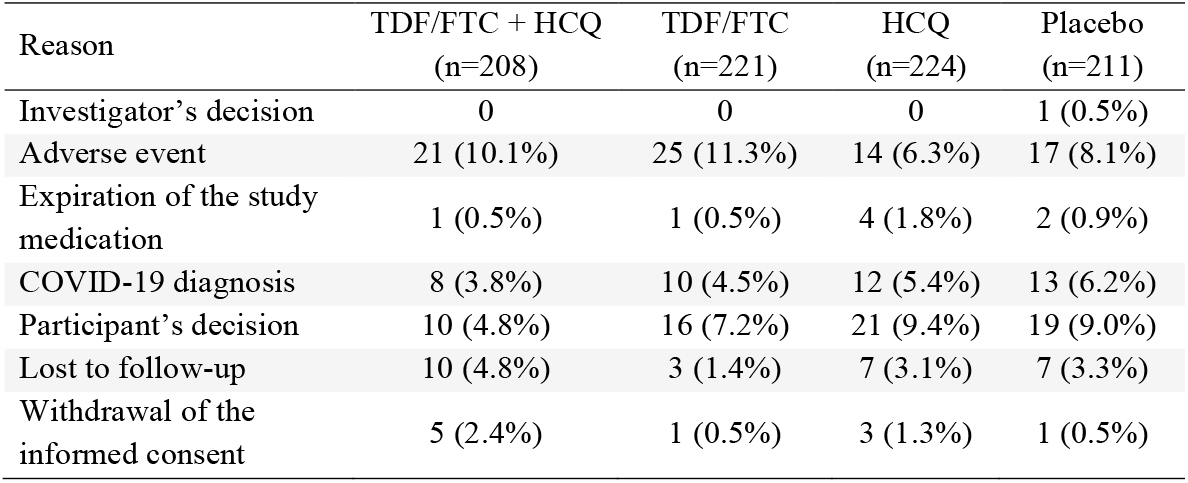
Reasons for early termination of the study medication by treatment group, EPICOS randomized trial.

| Reason | TDF/FTC + HCQ<br>(n=208) | TDF/FTC<br>(n=221) | HCQ<br>(n=224) | Placebo<br>(n=211) |
| --- | --- | --- | --- | --- |
| Investigator's decision | 0 | 0 | 0 | 1 (0.5%) |
| Adverse event | 21 (10.1%) | 25 (11.3%) | 14 (6.3%) | 17 (8.1%) |
| Expiration of the study medication | 1 (0.5%) | 1 (0.5%) | 4 (1.8%) | 2 (0.9%) |
| COVID-19 diagnosis | 8 (3.8%) | 10 (4.5%) | 12 (5.4%) | 13 (6.2%) |
| Participant's decision | 10 (4.8%) | 16 (7.2%) | 21 (9.4%) | 19 (9.0%) |
| Lost to follow-up | 10 (4.8%) | 3 (1.4%) | 7 (3.1%) | 7 (3.3%) |
| Withdrawal of the informed consent | 5 (2.4%) | 1 (0.5%) | 3 (1.3%) | 1 (0.5%) |

**Supplementary Table 2.** Reasons for incomplete follow-up by treatment group, EPICOS randomized trial.

| Reason | TDF/FTC + HCQ<br>(n=220) | TDF/FTC<br>(n=233) | HCQ<br>(n=231) | Placebo<br>(n=223) |
| --- | --- | --- | --- | --- |
| Investigator's decision | 0 | 0 | 1 (0.4%) | 2 (0.9%) |
| Adverse event | 9 (4.1%) | 9 (3.9%) | 7 (3.0%) | 7 (3.1%) |
| COVID-19 vaccine | 2 (0.9%) | 1 (0.4%) | 1 (0.4%) | 3 (1.3%) |
| Participant's decision | 12 (5.5%) | 17 (7.3%) | 19 (8.2%) | 17 (7.6%) |
| Lost to follow-up | 12 (5.5%) | 11 (4.7%) | 11 (4.8%) | 14 (6.3%) |
| Withdrawal of<br>informed consent | 5 (2.3%) | 2 (0.9%) | 4 (1.7%) | 2 (0.9%) |
| Unknown | 12 (5.5%) | 12 (5.2%) | 7 (3.0%) | 12 (5.4%) |

**Supplementary Table 3.**
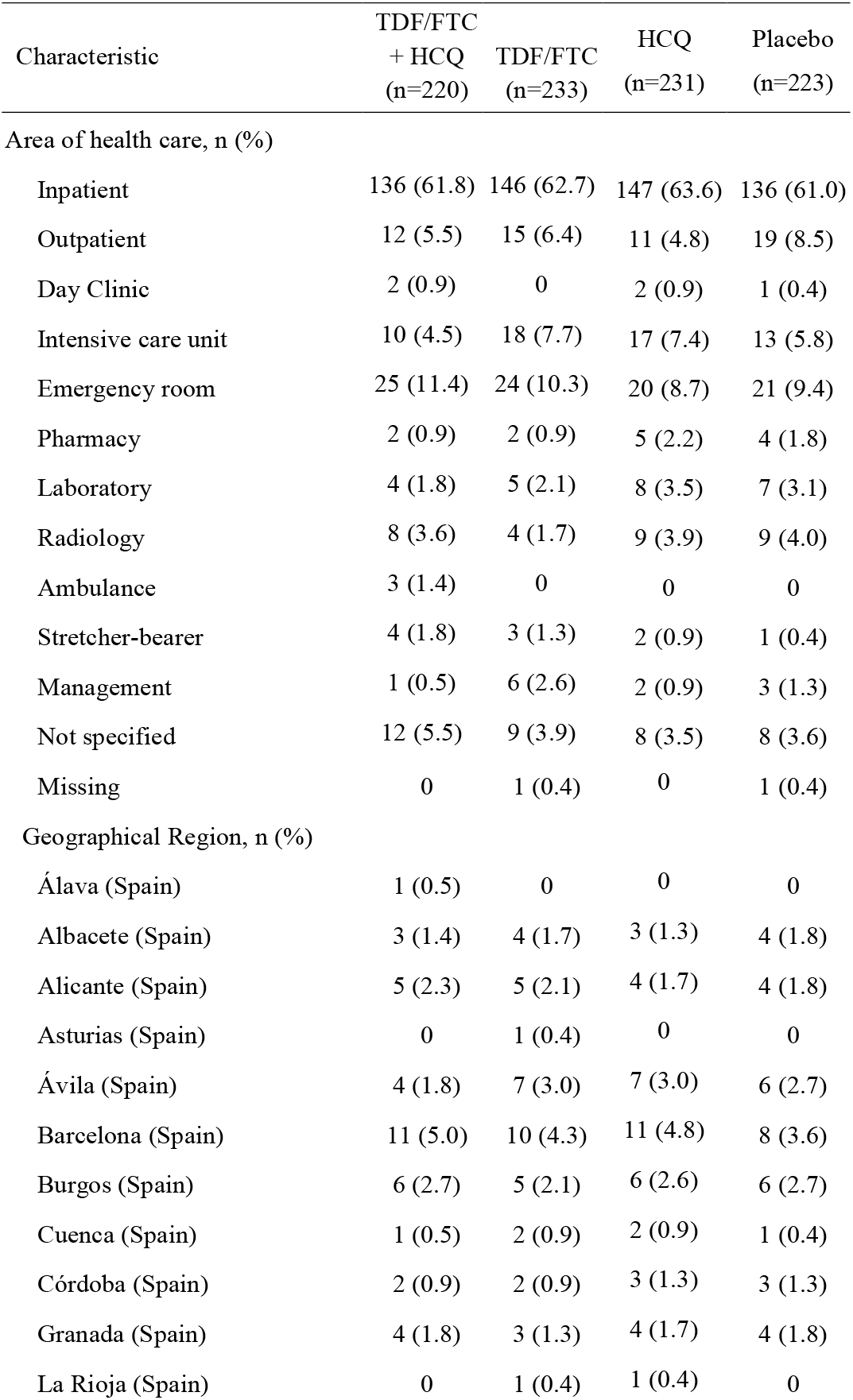
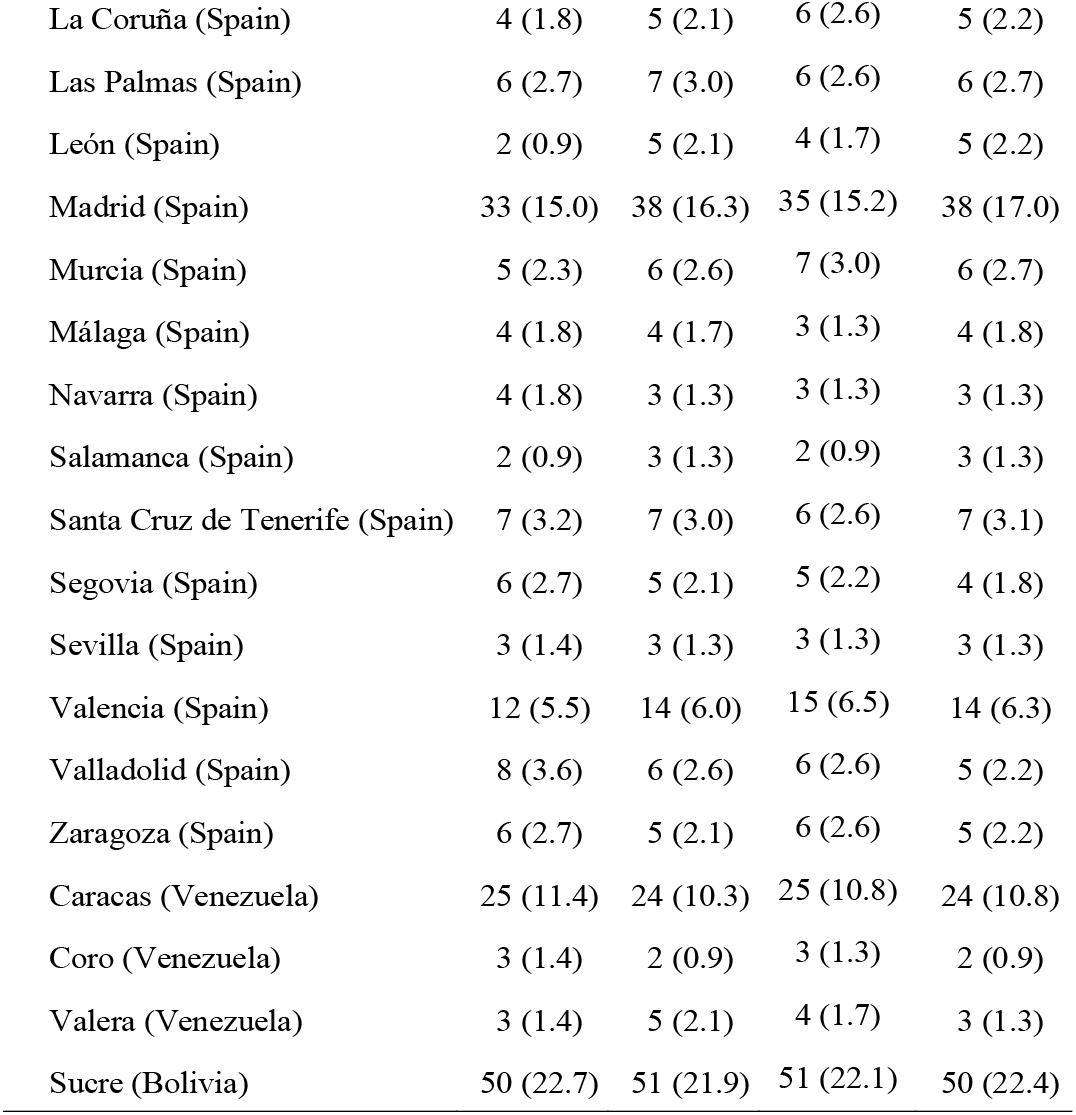
Supplementary baseline characteristics of 907 participants, EPICOS randomized trial

| Characteristic | TDF/FTC<br>+ HCQ<br>(n=220) | TDF/FTC<br>(n=233) | HCQ<br>(n=231) | Placebo<br>(n=223) |
| --- | --- | --- | --- | --- |
| Area of health care, n (%) |  |  |  |  |
| Inpatient | 136 (61.8) | 146 (62.7) | 147 (63.6) | 136 (61.0) |
| Outpatient | 12 (5.5) | 15 (6.4) | 11 (4.8) | 19 (8.5) |
| Day Clinic | 2 (0.9) | 0 | 2 (0.9) | 1 (0.4) |
| Intensive care unit | 10 (4.5) | 18 (7.7) | 17 (7.4) | 13 (5.8) |
| Emergency room | 25 (11.4) | 24 (10.3) | 20 (8.7) | 21 (9.4) |
| Pharmacy | 2 (0.9) | 2 (0.9) | 5 (2.2) | 4 (1.8) |
| Laboratory | 4 (1.8) | 5 (2.1) | 8 (3.5) | 7 (3.1) |
| Radiology | 8 (3.6) | 4 (1.7) | 9 (3.9) | 9 (4.0) |
| Ambulance | 3 (1.4) | 0 | 0 | 0 |
| Stretcher-bearer | 4 (1.8) | 3 (1.3) | 2 (0.9) | 1 (0.4) |
| Management | 1 (0.5) | 6 (2.6) | 2 (0.9) | 3 (1.3) |
| Not specified | 12 (5.5) | 9 (3.9) | 8 (3.5) | 8 (3.6) |
| Missing | 0 | 1 (0.4) | 0 | 1 (0.4) |
| Geographical Region, n (%) |  |  |  |  |
| Álava (Spain) | 1 (0.5) | 0 | 0 | 0 |
| Albacete (Spain) | 3 (1.4) | 4 (1.7) | 3 (1.3) | 4 (1.8) |
| Alicante (Spain) | 5 (2.3) | 5 (2.1) | 4 (1.7) | 4 (1.8) |
| Asturias (Spain) | 0 | 1 (0.4) | 0 | 0 |
| Ávila (Spain) | 4 (1.8) | 7 (3.0) | 7 (3.0) | 6 (2.7) |
| Barcelona (Spain) | 11 (5.0) | 10 (4.3) | 11 (4.8) | 8 (3.6) |
| Burgos (Spain) | 6 (2.7) | 5 (2.1) | 6 (2.6) | 6 (2.7) |
| Cuenca (Spain) | 1 (0.5) | 2 (0.9) | 2 (0.9) | 1 (0.4) |
| Córdoba (Spain) | 2 (0.9) | 2 (0.9) | 3 (1.3) | 3 (1.3) |
| Granada (Spain) | 4 (1.8) | 3 (1.3) | 4 (1.7) | 4 (1.8) |
| La Rioja (Spain) | 0 | 1 (0.4) | 1 (0.4) | 0 |
| La Coruña (Spain) | 4 (1.8) | 5 (2.1) | 6 (2.6) | 5 (2.2) |
| Las Palmas (Spain) | 6 (2.7) | 7 (3.0) | 6 (2.6) | 6 (2.7) |
| León (Spain) | 2 (0.9) | 5 (2.1) | 4 (1.7) | 5 (2.2) |
| Madrid (Spain) | 33 (15.0) | 38 (16.3) | 35 (15.2) | 38 (17.0) |
| Murcia (Spain) | 5 (2.3) | 6 (2.6) | 7 (3.0) | 6 (2.7) |
| Málaga (Spain) | 4 (1.8) | 4 (1.7) | 3 (1.3) | 4 (1.8) |
| Navarra (Spain) | 4 (1.8) | 3 (1.3) | 3 (1.3) | 3 (1.3) |
| Salamanca (Spain) | 2 (0.9) | 3 (1.3) | 2 (0.9) | 3 (1.3) |
| Santa Cruz de Tenerife (Spain) | 7 (3.2) | 7 (3.0) | 6 (2.6) | 7 (3.1) |
| Segovia (Spain) | 6 (2.7) | 5 (2.1) | 5 (2.2) | 4 (1.8) |
| Sevilla (Spain) | 3 (1.4) | 3 (1.3) | 3 (1.3) | 3 (1.3) |
| Valencia (Spain) | 12 (5.5) | 14 (6.0) | 15 (6.5) | 14 (6.3) |
| Valladolid (Spain) | 8 (3.6) | 6 (2.6) | 6 (2.6) | 5 (2.2) |
| Zaragoza (Spain) | 6 (2.7) | 5 (2.1) | 6 (2.6) | 5 (2.2) |
| Caracas (Venezuela) | 25 (11.4) | 24 (10.3) | 25 (10.8) | 24 (10.8) |
| Coro (Venezuela) | 3 (1.4) | 2 (0.9) | 3 (1.3) | 2 (0.9) |
| Valera (Venezuela) | 3 (1.4) | 5 (2.1) | 4 (1.7) | 3 (1.3) |
| Sucre (Bolivia) | 50 (22.7) | 51 (21.9) | 51 (22.1) | 50 (22.4) |

**Supplementary Table 4.** Frequency of symptoms accompanying SARS-CoV-2 infections, EPICOS randomized trial

| Reason | TDF/FTC + HCQ<br>(n=13) | TDF/FTC<br>(n=20) | HCQ<br>(n=21) | Placebo<br>(n=23) |
| --- | --- | --- | --- | --- |
| Fever | 2 (15.4%) | 2 (10.0%) | 1 (4.8%) | 4 (17.4%) |
| Cough | 0 | 0 | 2 (9.5%) | 1 (4.3%) |
| Shortness of breath | 1 (7.7%) | 1 (5.0%) | 0 | 0 |
| Respiratory insufficiency | 0 | 0 | 0 | 0 |
| Malaise | 3 (23.1%) | 2 (10.0%) | 2 (9.5%) | 3 (13.0%) |
| Arthromyalgia | 2 (15.4%) | 0 | 2 (9.5%) | 2 (8.7%) |
| Asthenia | 1 (7.7%) | 1 (4.5%) | 1 (4.8%) | 1 (4.3%) |
| Headache | 3 (23.1%) | 3 (15.0%) | 2 (9.5%) | 3 (13.0%) |
| Vomiting | 0 | 0 | 1 (4.8%) |  |
| Diarrhea | 0 | 0 | 1 (4.8%) | 1 (4.3%) |
| Olfactory loss | 1 (7.7%) | 1 (5.0%) | 0 | 0 |
| Taste loss | 1 (7.7%) | 1 (5.0%) | 0 | 0 |
| Hospital admission | 0 | 0 | 0 | 0 |

**Supplementary Table 5.** Estimated 14-week risks of symptomatic, asymptomatic, and any COVID-19 diagnosis in HCQ-containing groups and groups without HCQ, EPICOS randomized trial.

| **Symptomatic COVID-19** | Cases / n | 14-week risk<br>(95% CI), % | Risk difference<br>(95% CI), % | Risk ratio<br>(95% CI) |
| --- | --- | --- | --- | --- |
| HCQ-containing groups | 6 / 451 | 1.25 (0.23 to 2.31) | -0.58 (-2.39 to 1.02) | 0.68 (0.10 to 2.04) |
| Groups without HCQ | 8 / 456 | 1.82 (0.72 to 3.21) | Reference | Reference |
| **Asymptomatic COVID-19** |  |  |  |  |
| HCQ-containing groups | 28 / 451 | 7.33 (4.61 to 10.22) | -1.93 (-5.82 to 2.23) | 0.79 (0.47 to 1.33) |
| Groups without HCQ | 35 / 456 | 9.25 (6.22 to 12.15) | Reference | Reference |
| **Any COVID-19** |  |  |  |  |
| HCQ-containing groups | 34 / 451 | 8.53 (5.61 to 11.71) | -2.43 (-6.56 to 1.95) | 0.78 (0.49 to 1.23) |
| Groups without HCQ | 43 / 456 | 10.96 (8.05 to 14.03) | Reference | Reference |

CI: confidence interval

**Supplementary Table 6.** Estimated 14-week risks of symptomatic, asymptomatic, and any COVID-19 diagnosis in TDF/FTC-containing groups and groups without TDF/FTC, EPICOS randomized trial.

| **Symptomatic COVID-19** | Cases / n | 14-week risk<br>(95% CI), % | Risk difference<br>(95% CI), % | Risk ratio<br>(95% CI) |
| --- | --- | --- | --- | --- |
| TDF-containing groups | 6 / 453 | 1.02 (0.24 to 2.16) | -1.05 (-2.69 to 0.65) | 0.49 (0.09 to 1.70) |
| Groups without TDF | 8 / 454 | 2.07 (0.78 to 3.36) | Reference | Reference |
| **Asymptomatic COVID-19** |  |  |  |  |
| TDF-containing groups | 27 / 453 | 7.05 (4.24 to 9.57) | -2.50 (-6.68 to 1.43) | 0.74 (0.43 to 1.21) |
| Groups without TDF | 36 / 454 | 9.55 (6.43 to 12.41) | Reference | Reference |
| **Any COVID-19** |  |  |  |  |
| TDF-containing groups | 33 / 453 | 8.01 (5.33 to 10.93) | -3.51 (-7.56 to 0.81) | 0.70 (0.43 to 1.10) |
| Groups without TDF | 44 / 454 | 11.52 (7.94 to 14.63) | Reference | Reference |

CI: confidence interval

**Supplementary Table 7.** Estimated hazard ratios of symptomatic, asymptomatic, and any COVID-19 diagnosis by treatment group, EPICOS randomized trial

| **Symptomatic COVID-19** | **Hazard ratio**<br>**(95% CI)** |
| --- | --- |
| TDF/FTC + HCQ | 0.40 (0.08 to 2.06) |
| TDF/FTC | 0.37 (0.07 to 1.91) |
| HCQ | 0.56 (0.13 to 2.32) |
| Placebo | Reference |
| HCQ groups | 0.71 (0.23 to 2.25) |
| HCQ placebo groups | Reference |
| TDF groups | 0.50 (0.15 to 1.66) |
| TDF placebo groups | Reference |
| **Asymptomatic COVID-19** |  |
| TDF/FTC + HCQ | 0.55 (0.25 to 1.18) |
| TDF/FTC | 0.81 (0.41 to 1.58) |
| HCQ | 0.93 (0.49 to 1.79) |
| Placebo | Reference |
| HCQ groups | 0.83 (0.50 to 1.37) |
| HCQ placebo groups | Reference |
| TDF groups | 0.71 (0.43 to 1.17) |
| TDF placebo groups | Reference |
| **Any COVID-19** |  |
| TDF/FTC + HCQ | 0.51 (0.26 to 1.03) |
| TDF/FTC | 0.71 (0.38 to 1.32) |
| HCQ | 0.85 (0.47 to 1.54) |
| Placebo | Reference |
| HCQ groups | 0.81 (0.51 to 1.28) |
| HCQ placebo groups | Reference |
| TDF groups | 0.67 (0.42 to 1.06) |
| TDF placebo groups | Reference |

**Supplementary Figure 1.**
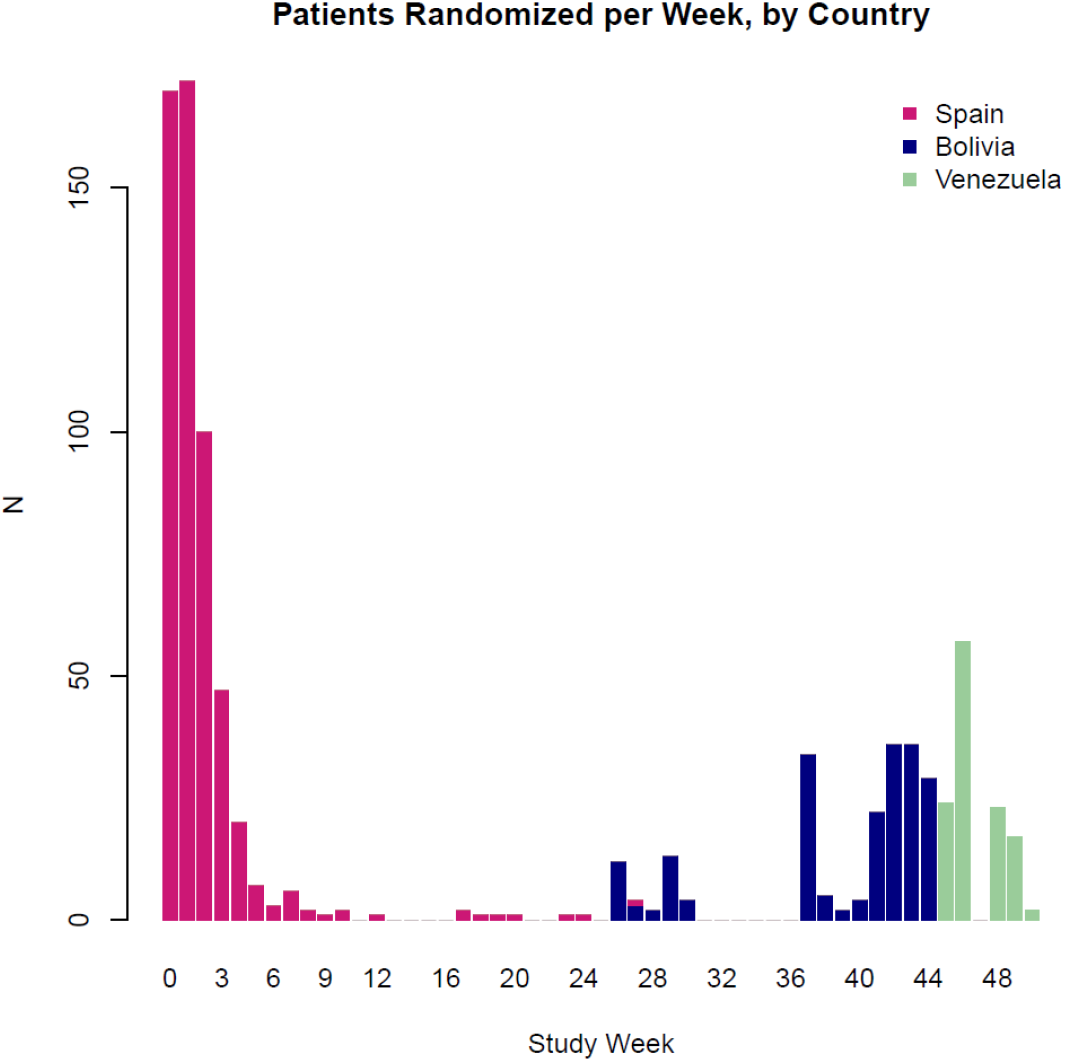
Recruitment by study week and by country, EPICOS randomized trial.

**Supplementary Figure 2.**
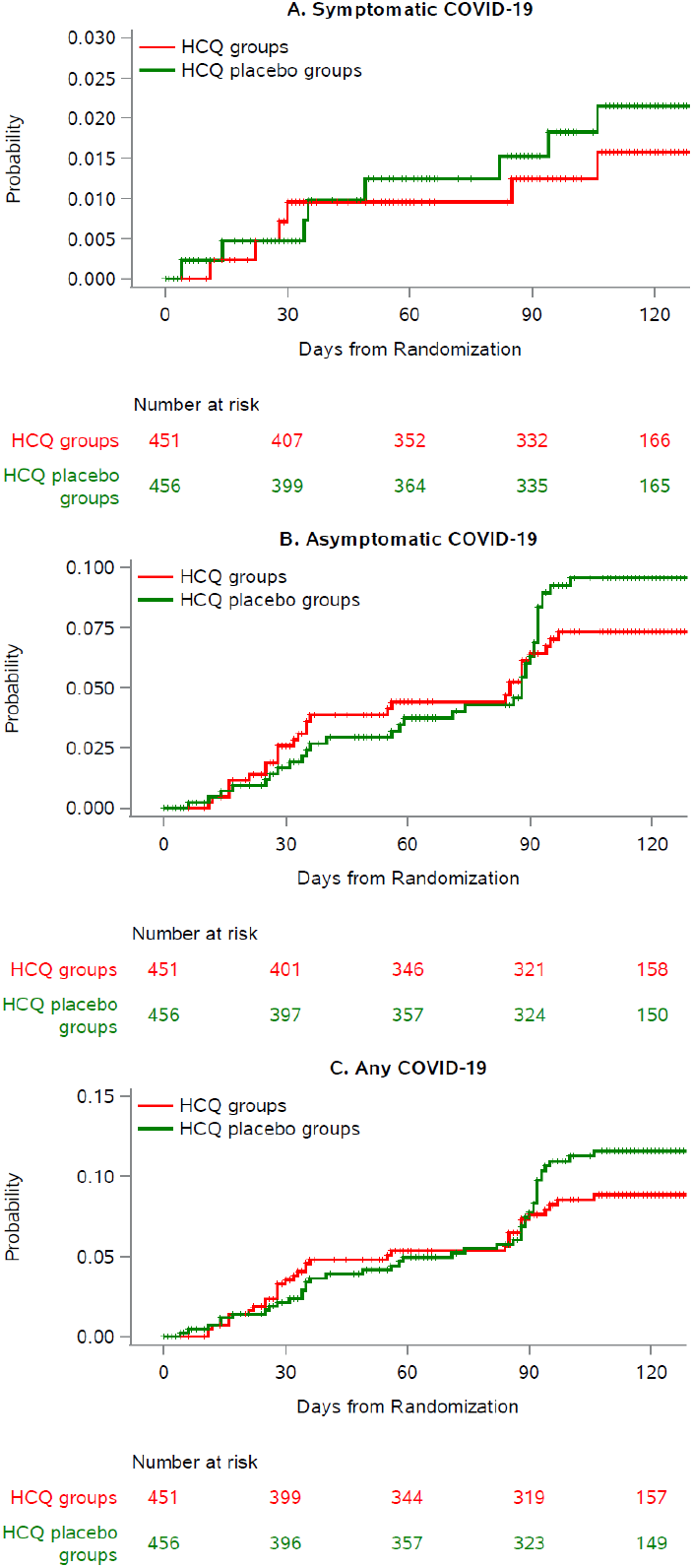
Cumulative risk of symptomatic and asymptomatic COVID-19 HCQ-containing groups and groups without HCQ, EPICOS randomized trial

**Supplementary Figure 3.**
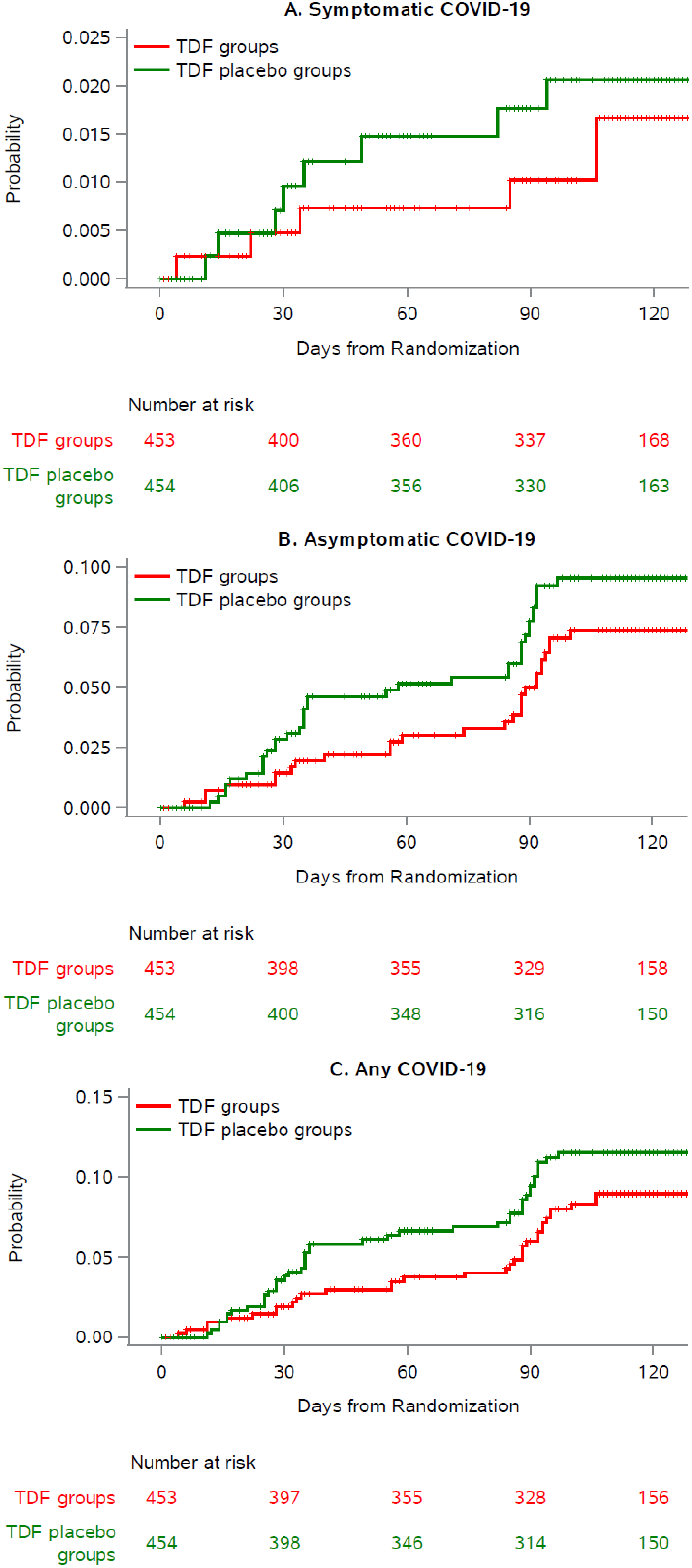
Cumulative risk of symptomatic and asymptomatic COVID-19 in TDF/FTC-containing groups and groups without TDF/FTC, EPICOS randomized trial.

